# Measurement of multiple SARS-CoV-2 antibody titer after vaccination represents individual vaccine response and contributes to individually appropriate vaccination schedules

**DOI:** 10.1101/2021.05.21.21257575

**Authors:** Kenji Ota, Satoshi Murakami, Hiroshi Mukae, Shigeru Kohno, Katsunori Yanagihara

## Abstract

The SARS-CoV-2 mRNA vaccine BNT162b2 (Pfizer-BioNTech) has become a game-changer in the COVID-19 crisis. Faster and wider coverage of vaccine supply is urgently needed, although vaccine supply is far from abundant in many countries and regions. There is no denying that Japan is vaccinating at a slower pace than in many other countries. At the end of April 2021, it had administered less than 2% of the population, according to the statistics website Our World in Data.^1^ Single-dose vaccination may be considered to induce an adequate antibody response in individuals with prior infection,^2–5^ which can lead to more effective vaccine distribution to people in serious need. However, multiple antibody responses in populations with various backgrounds, including prior COVID-19 infection, underlying diseases, and age, have not been investigated sufficiently. Due to the lack of antibody measurement data, antibody follow-up measurement recommendations have yet to be suggested. Accumulation of available data regarding quantitative antibody titer will lead to the establishment of personally customized schedules, including antibody follow-up, and will balance both faster delivery of vaccines and confirmation of effective vaccination.

---

We have been conducting a prospective study to evaluate antibody response after vaccination in healthcare workers with (n=23) and without (n=113) prior COVID-19 infection (confirmed by molecular testing) by measuring immunoglobulin (Ig) G and IgM against the SARS-CoV-2 receptor-binding domain (RBD) and IgG against the SARS-CoV-2 nucleocapsid (N) (Alinity, Abbott Laboratories, Chicago, IL). IgG (RBD) and IgM (RBD) are expected to be induced by both prior infection and vaccination, and IgG (N) only by prior infection. The measurement of IgG (RBD) by Alinity is reportedly well-correlated with the neutralization assay. This study was approved by the institutional review board of the Nagasaki University Hospital (approval number 21030401-2).

Baseline antibody profiles of the participants with prior infection indicated that IgG (RBD) peaked after 12-24 weeks of natural infection, IgM (RBD) showed a consistent decrease, and IgG (N) significantly decreased at more than 24 weeks of past infection (figure A). As concordant with previous reports^2–5^, single-dose vaccination in participants with prior infection yielded comparable or higher IgG (RBD) response to two-dose vaccination in participants without prior infection (mean ± standard deviation, 31523 ± 14,332 arbitrary units [AU] per mL vs. 22,461 ± 15,661 AU/mL, *P* = 0·01, figure B). IgM (RBD) response was observed in participants without prior infection at 14 days after the first vaccination, achieving a comparable antibody titer compared with those with prior infection (1·41 ± 1·93 chemiluminescence of Sample / Calibrator [S/C] vs. 1·96 ± 2·49 S/C, *P* = 0·24, figure C). However, no increase in IgM (RBD) was observed after the first or second dose of the vaccine in those with prior infection. IgG (N) showed no reaction by vaccination, concordant with our hypothesis, suggesting its usefulness in distinguishing prior history of infection (figure D). Only three of 113 uninfected participants showed IgG (RBD) below the cut-off level (50 AU/mL) at 14 days after vaccination; one of them had rheumatoid arthritis, and the other had collagen disease, under treatment by prednisolone, tacrolimus, and mycophenolate mofetil. The latter did not achieved immunization even at 7 days after 2^nd^ vaccination. From the viewpoint of the relationship between age and antibody response, age was inversely correlated with IgG (RBD) antibody response. We divided participants without prior infection into two groups (≥40 years and <40 years) to assess the differences in antibody response according to age (figure E). IgG (RBD) response was significantly lower in those >40 years old (19,087 ± 14,630 AU/mL vs. 25,334 ± 15,849 AU/mL, *P* = 0·04).

**Figure:**
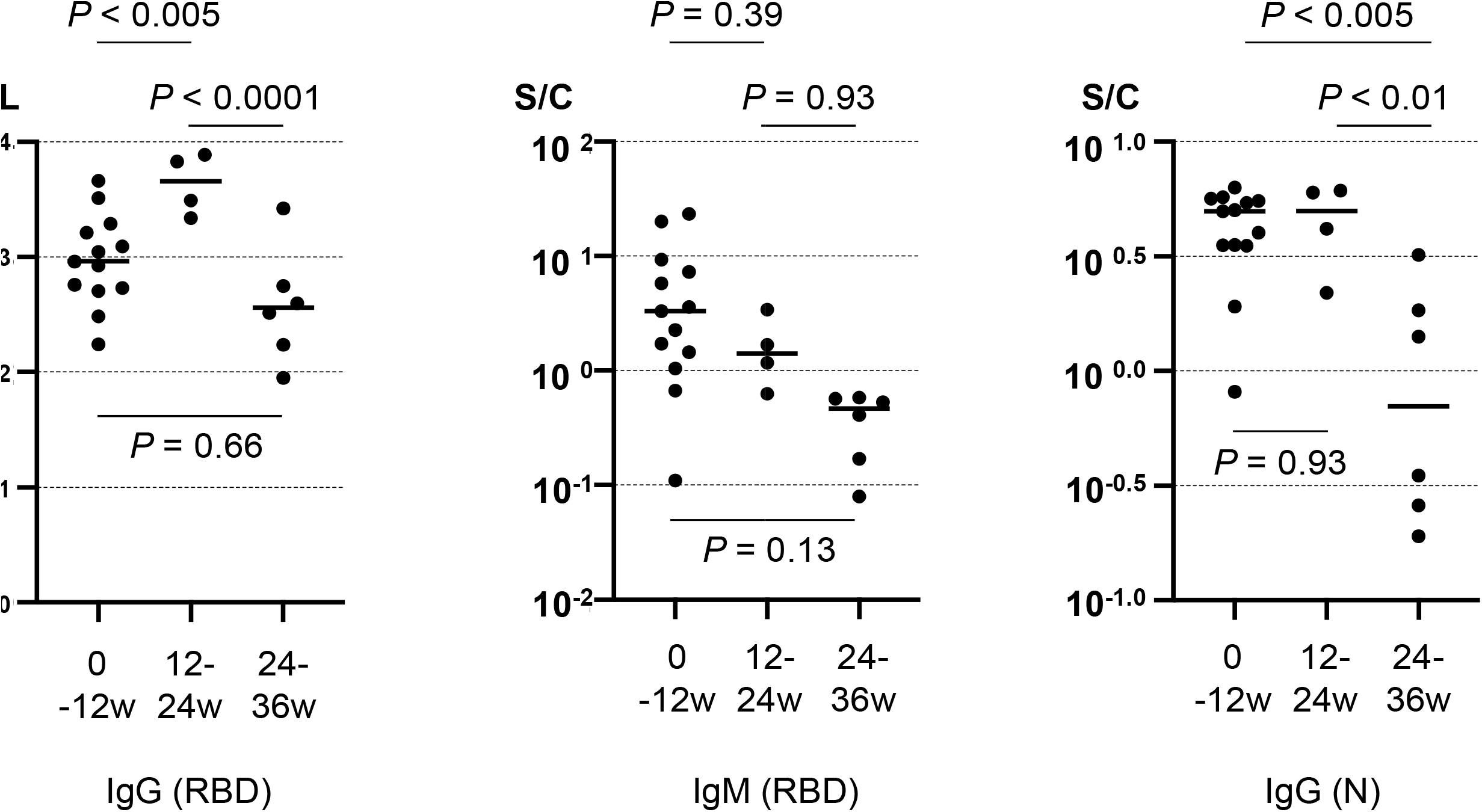
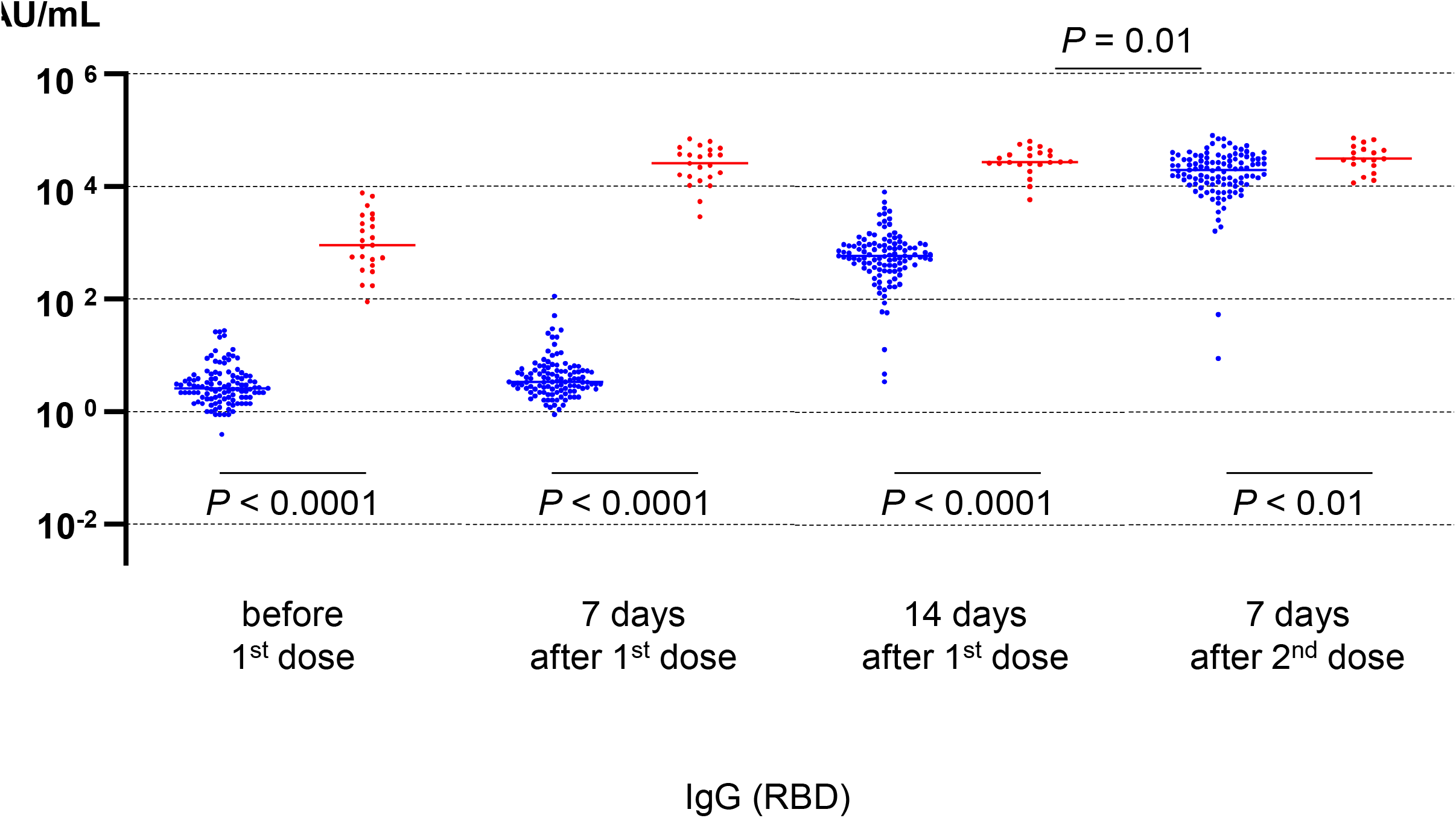
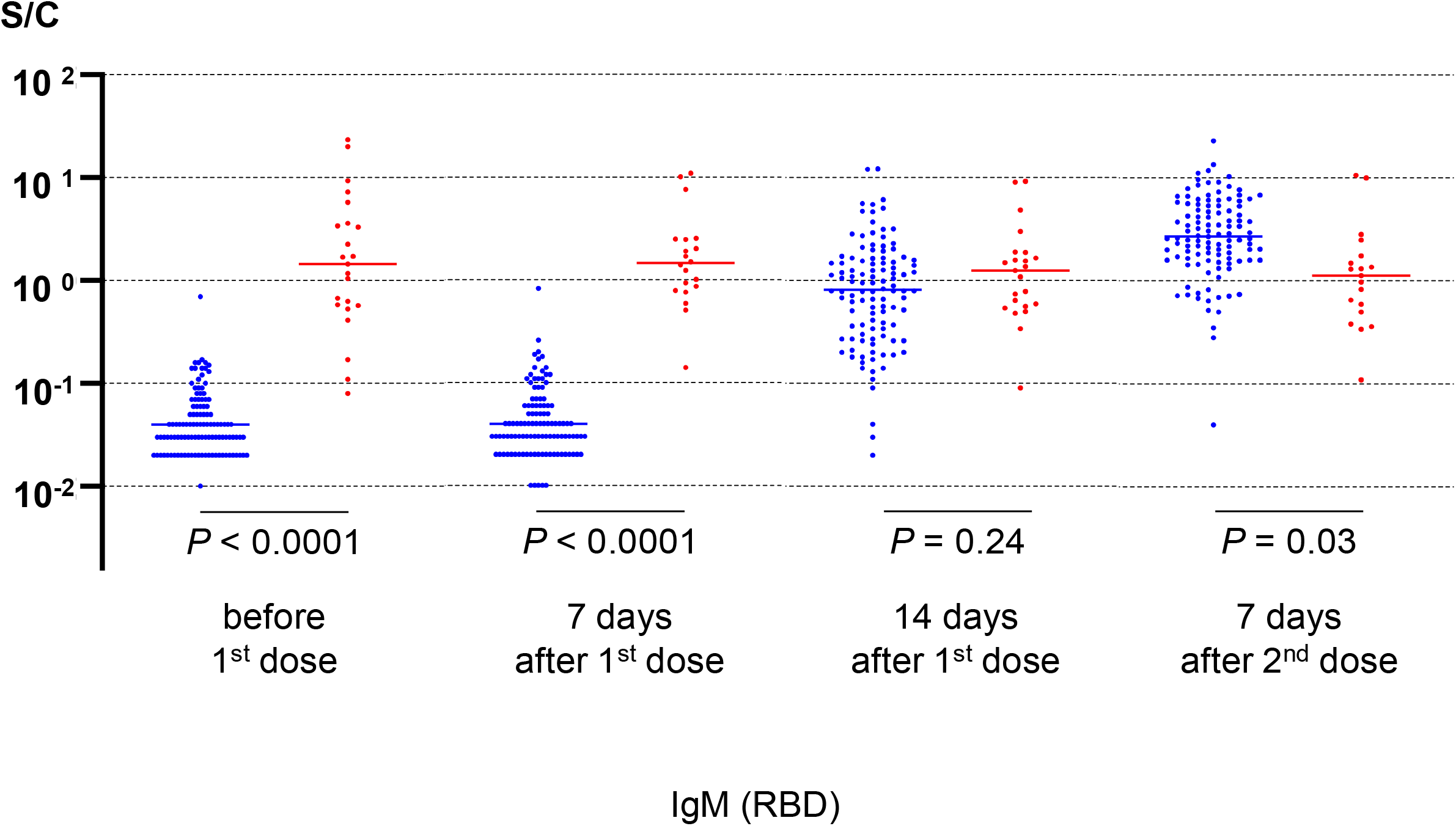
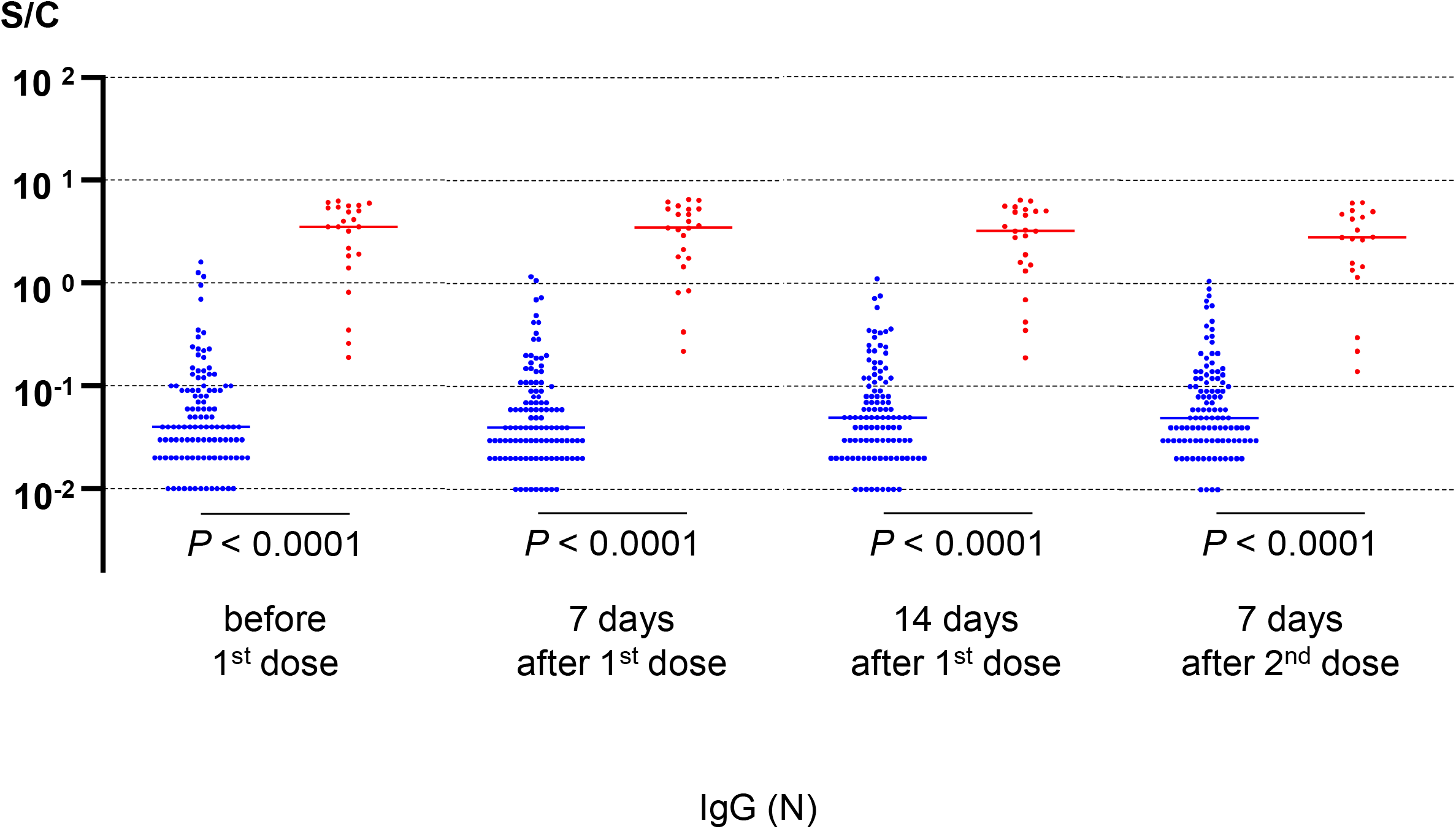
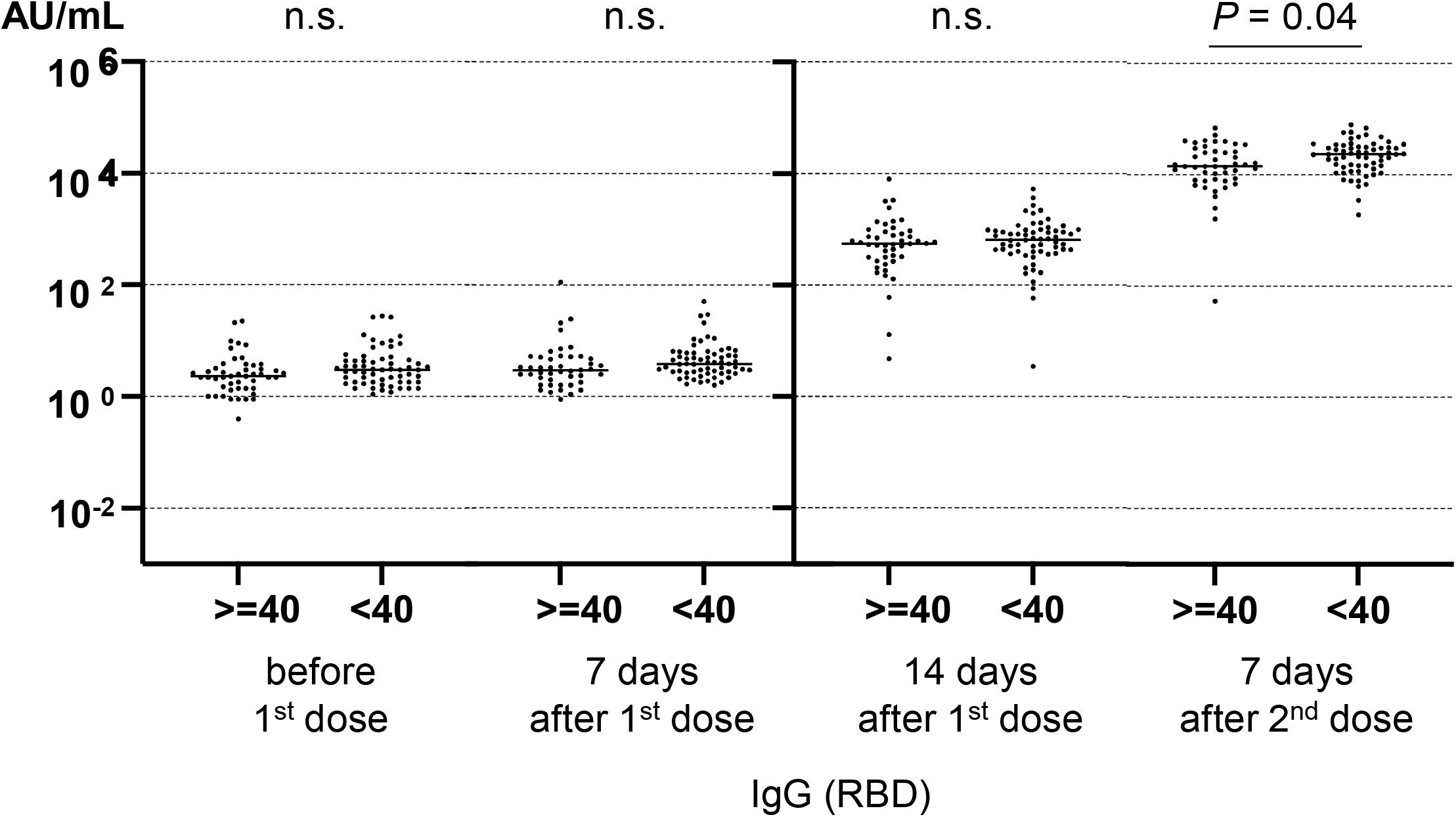
Multiple antibody titer in individuals before and after vaccination. (A) IgG (RBD), IgM (RBD), and IgG (N) in individuals with prior COVID-19 infections are shown. They were divided into three groups (0-12 weeks n=14, 12-24 weeks n=3, 24-36 weeks n=6) according to the time from infection to antibody measurement before vaccine. We applied Tukey’s multiple comparison test to assess the differences between groups. RBD, receptor-binding domain; N, nucleocapsid. S/C is index value, calculated according chemiluminescence of Sample / mean chemiluminescence of Calibrator. (B) (C) (D) IgG (RBD), IgM (RBD), and IgG (N) in individuals without and with prior COVID-19 infections are shown. Blue dots represent antibody titers of individuals without prior infection, and red dots represent prior infection. All participants analyzed in this report had two vaccinations three weeks apart. Each antibody was measured at four timepoints (7 days before vaccination, and 7 and 14 days after 1^st^ vaccination, and 7 days after 2^nd^ vaccination; ±1 days was accepted, sample number of individuals without and with prior infection 113 vs. 23, 111 vs. 23, 110 vs. 23, and 111 vs. 19, respectively). We applied a t-test to assess the differences between groups. (E) IgG (RBD) in individuals without prior COVID-19 infections are shown. They were divided into two groups (≥40 years old and <40 years old, n=47 and 63, respectively, one low responder was excluded from <40 years old group) according to the age at vaccination. We applied a t-test to assess the differences between groups.

According to our results, personal customized vaccination schedules together with appropriate follow-up may be proposed as follows.

Single-dose vaccination may be warranted for those with prior COVID-19 infection if an adequate antibody titer is confirmed. For persons with underlying diseases (e.g., rheumatoid disease, collagen diseases, or hemodialysis), follow-up by antibody measurement may be considered. Age in the forties can be considered a predictive factor for lower antibody response.

In addition, this observational study suggests that when evaluating antibody responses induced by vaccines, measuring IgG (RBD) is more useful than other antibodies, since IgG (RBD) showed evident response.

The compatibility of both speedy vaccine delivery and effective vaccination schedules is our top concern in conquering COVID-19. We must pay close attention to adapting evidence to clinical practice, since it may unexpectedly lead to worse outcomes of ineffective vaccination. Therefore, our argument is that monitoring each person’s antibody titer is warranted in public with expected low and high responders. Further continuous observation (up to 48 weeks) is in progress to confirm the duration of the antibody response obtained by vaccination.

## Data Availability

The data that support the findings of this study are available from the corresponding author, KY, upon reasonable request.

